# Senior dental students reflective activities involving community-service learning

**DOI:** 10.64898/2025.12.21.25342789

**Authors:** Mario Brondani, Bebecca Elias, Renata Paz Leal Pereira

## Abstract

**Objective:** Community service-learning (CSL) placements engage with equity-deserving groups. They receive oral health care and students can critically reflect on their experiences. This study aimed to thematically explore the reflections of senior dental students providing oral health services to equity-deserving communities in British Columbia, Canada.

**Methods:** Semi-structured written reflections were collected cross-sectionally from three consecutive graduating cohorts of fourth-year dental students (2022–23, 2023–24, and 2024–25). Reflections were a mandatory component of a community placement course, were approximately 500 words in length, and were prompted as follows: “Describe your personal experience at the assigned community clinic, noting moments of revelation, valuable learning, and/or disappointment for you.” An exploratory thematic analysis was conducted using an iterative coding process to identify and interpret categories and themes.

**Results:** From all the three years, 764 reflections were collected (191-625 words each) from 171 students. Of these, 124 reflections were excluded because they consisted solely of descriptions of procedures. Data saturation was reached after in-depth analysis of 205 reflections, yielding four overarching themes, including ‘learning across differences’; and ‘pause-breathe-refine’. These themes were informed by categories highlighting that detrimental impact of overly controlling mentorship styles and observation-only experiences on students’ learning.

**Conclusion:** Transformative experiences were observed, while students also reflected on less positive practices. Students emphasized the importance of CSL placements for their education, professional growth, and understanding of underserved populations, while also highlighting implementation challenges. Future research should examine the long-term impact of CSL activities once these challenges are addressed.

## Introduction

In contemporary dental curricula, community service-learning (CSL) has become a central feature by combining clinical care for equity-deserving communities with opportunities for students to reflect on whom they serve and how they serve them.^1,2^ A substantial body of evidence demonstrates the educational value of CSL and community-based clinical experiences. Attitudinally, service-learning seems to be associated with stronger commitments to community engagement and an increased intention to treat equity-seeking populations after graduation.^3,4^ Clinically, students report (and faculty observe) independence, efficiency, and adaptability in lower-resource, higher-throughput settings. These placements also offer broader exposure to preventive, operative, and community-tailored procedures than traditional university dental clinics.^5,6,7^ Importantly, CSL experiences are both educationally beneficial and service-producing, addressing unmet population needs while diversifying students’ clinical experience.^6^

CSL activities offer learning gains that are further enhanced when students engage in structured reflection about such experiences. Grounded in experiential and reflective learning theories,^1^ CSL encourages students to connect clinical encounters with broader social determinants of oral health and with their emerging professional identities; reflections should go beyond a mere description of events.^8^ In dentistry, reflective exercises have been shown to deepen self-awareness, challenge assumptions about patients’ lives, and improve communication, empathy, and confidence. These are skills that are difficult to cultivate through simulation or didactic instruction alone.^8,9,10^ Structured or semi-structured reflections, guided by targeted prompts, help students link clinical decisions to patients’ socioeconomic contexts, recognize personal biases, and articulate ethical and social responsibilities, even in resource-constrained environments.^9,11,12^

But even with reflections aside, the implementation of CSL is not without challenges. These include variable supervision and assessment standards across community sites, logistical burdens (e.g., travel, scheduling, documentation), and sustainability pressures for host clinics.^13^ There is also academic reluctance to send students outside the controlled environment of university clinics, which offer abundant faculty oversight, standardized equipment, and predictable workflows.^5^ Without adequate preparation and debriefing, students may experience anxiety when encountering unfamiliar procedures, language barriers, or patients with complex psychosocial needs. These challenges underscore the importance of intentionally designed CSL programs with clear learning objectives, preparatory training, equitable community partnerships, and structured reflective components to maximize benefits for both learners and patients.^14^ Nonetheless, the existing literature remains limited in examining whether these challenges, or additional ones, surface when students are asked to reflect about the CSL activity. Also limited is the knowledge around the impact of such challenges as experienced by students. These gaps underscore the need for research that foregrounds students’ perspectives.

At the Faculty of Dentistry of the University of British Columbia, reflective exercises are embedded across the curriculum, from preclinical courses in junior years to CSL experiences in senior years that complement, but do not replicate, university clinic training.^2,5,6,9,11^ In the fourth and final year of undergraduate dental education, students complete semi-structured reflections guided by a prompt designed to foster critical appraisal of clinical experiences across four different community clinics; students also submit reflections as part of other activities, as we have previously studied.^2,9,10,18^ Yet, these reflections in particular had not previously been analyzed beyond course requirements. This study therefore examined fourth-year dental students’ reflections generated during CSL placements delivering oral health care at equity-deserving populations in British Columbia. Specifically, we employ a qualitative methodology to thematically explore moments of revelation and struggle experienced by senior dental students as they reflected on providing oral health care in community settings as part of their dental educational training.

## Methods

The University of British Columbia’s Behavioural Research Ethics Board approved this study (H25-01322). Data consisted of semi-structured written reflections submitted by three consecutive cohorts of graduating 4^th^ year dental students (2022–23, 2023–24, and 2024–25). A total of 764 reflections were collected: 330 from 55 students (2022–23), 236 from 59 students (2023–24), and 198 from 57 students (2024–25). A 500-word reflection was submitted by students multiple time, prompted by: “Describe your personal experience at the assigned community clinic, noting moments of revelation, valuable learning, and/or disappointment for you.” Reflections were intended to encourage consideration of unfamiliar or thought-provoking experiences distinct from those encountered in the university dental clinic. Students were explicitly asked to move beyond listing procedures and treatments provided. Reflections were mandatory (pass, if completed; fail, if not completed) but not graded and were submitted electronically through the Canvas® learning management system. Each reflection corresponded to a full-day experience at one of four community sites, including:

Community site A: A not-for-profit dental clinic in Vancouver’s Downtown Eastside providing affordable general dentistry primarily low-income residents, many of whom experience housing insecurity and substance use disorders.

Community site B: A large not-for-profit dental clinic within a medical center in East Vancouver offering integrated primary care and pharmaceutical services to patients with and without dental insurance.

Community site C: A hospital-based dental clinic focused on pediatric patients in Vancouver treated under general anesthesia; student participation was observational in nature.

Community site D: A non-profit community-based organization in the Lower Mainland providing integrated health and social services to women, girls, and children at risk or involved in the justice system.

The differences in the number of reflections across cohorts reflected changes in course structure and community partnerships: in 2022–23, students visited all four sites for a total of six visits each, with one reflection per visit; in 2023–24, students also visited four sites but for a total of four visits each, again submitting one reflection per visit; in 2024–25, students visited only two sites for a total of three visits each, with one reflection per visit.

### Thematic analysis

The aim of the thematic analysis is to understand meanings, concepts, and experiences portrayed within the reflections; it starts by exploring meanings and relationships of words, themes, or concepts.^16^ Codes were used to identify ideas in the form of a word or words in the reflections. These words—the codes—were grouped into categories representing related concepts, which were then synthesized into overarching themes.^17,15^ Four themes and their associated categories are presented in this manuscript; these are illustrative rather than exhaustive.

Of the 764 reflections, 124 (52 in 2022–23, 40 in 2023–24, and 32 in 2024–25) were excluded because they consisted solely of procedural descriptions. The remaining 640 reflections were securely downloaded from the Canvas® platform to the first author’s (MB) university computer. All reflections were anonymized by removing student names and identification numbers; only the academic year, student gender, and clinic site were retained for contextualization. De-identified reflections were printed and subjected to an inductive thematic analysis.^16^

As we have done previously,^9,17,18^ three identical sets of 10 randomly selected reflections were distributed to the authors (MB, RPLP, RE). The authors read them independently to develop a preliminary coding framework, and met to discuss the coding framework for consensus. The remaining 630 reflections were then analyzed independently by the authors (each received 210 different reflections each), with iterative refinement of codes, categories, and themes. Regular consensus meetings (between July 10 and August 20, 2025) ensured consistency and minimized interpretive bias.^19,20^ Data saturation was achieved after in-depth analysis of 180 reflections and confirmed through review of an additional 25 reflections, yielding a total of 205 reflections analyzed to saturation.

## Results

Four main themes and nine associated categories were identified:

- Impact of social determinants on access to oral health care, with the following categories: ‘not lost in translation’ and ‘financial abyss’;
- Professional growth, with the following categories: ‘working independently’, ‘collaborative practice’, and ‘enactive learning’;
- In spite of differences, with the following categories: ‘operational procedures’ and constrained resources’;
- Pause–breathe–refine, with the following categories: ‘vicarious quandary’, and foreigner environment’.

Across a large number of reflections, the theme **Impact of the Social Determinants on Access to Oral Health Care** emerged clearly. One illustrative observation came from a female student in 2024 assigned to Clinic C:

> “We were given the opportunity to see care being provided while considering the social determinants of health for each patient—an opportunity we would not have had by working solely at the university clinic.”

This theme encompassed several categories, including ‘Not Lost in Translation’, where students reflected on the importance of establishing clear communication with patients who have limited English proficiency:

> “Today my first patient spoke limited English, which made communication challenging. Much of the interaction relied on using an app to translate in order to obtain consent, inform the patient about the treatment, and provide post-operative instructions. Initially, it was difficult to determine whether the correct message was being conveyed. To ensure the patient understood what was happening, I also made drawings of the tooth and the procedure. This helped the process a lot.” (Male student, Clinic B, 2023)

> “The patient I saw did not speak English, and we had to use some creativity, such as Google Translate, to ensure she properly understood what we were looking for, which radiographs needed to be taken, and what treatment was recommended. We also allowed her to express her questions and concerns back to us. This emphasized for me the importance of accessibility to quality dental care.” (Female student, Clinic D, 2022)

> “The patient who required extensive restoration had a language barrier, and explaining procedures became challenging. I spoke slowly and used simple words, but since I was unsure whether she fully understood, we called her daughter so she could translate. Including her daughter in the treatment planning was very helpful.” (Male student, Clinic B, 2022)

The cost of care was another recurrent issue, categorized as ‘Financial Abyss’, reflecting the deep and often unbridgeable gap created by the high cost of oral health care:

> “What struck me most wasn’t just the clinical work itself, but the patient population and their complex needs. It was eye-opening to realize that for some, routine dental care had been a luxury they simply couldn’t afford, leading to extensive damage that is now difficult to repair and requires hospital care.” (Female student, Clinic C, 2025)

> “One of the most challenging aspects of this rotation was navigating the financial limitations patients face. While we presented all available treatment options, some patients chose temporary fixes, such as open-and-drain procedures, over more permanent solutions like root canals and crowns because they could not afford them.” (Female student, Clinic A, 2024)

> “This patient allowed me to see how socioeconomic barriers delay dental treatment. It highlighted the necessity of low-cost clinics and a public dental care plan that could help alleviate the financial burden of dental treatment for many.” (Male student, Clinic B, 2023)

The theme of **Professional Growth**, had three categories. ‘Working Independently’ emerged as a prominent category. Many students recognized their proximity to graduation and reflected on the confidence gained through these rotations:

> “As I prepare to graduate, I recognize that I’ll soon be expected to work independently, and I want to feel confident managing a procedure from beginning to end, as I had the opportunity to do here.” (Male student, Clinic B, 2024)

> “The overseeing dentist gave us the opportunity to make decisions and work independently rather than blindly following instructions. Although intimidating at times, it was great practice for us as soon-to-be graduates.” (Female student, Clinic A, 2022)

> “The most notable experience today was performing a surgical extraction without direct supervision, unlike at UBC. With graduation only a month away, this opportunity was very welcome. I did sweat a bit when the palatal root with a metal post fractured early on, but learning to troubleshoot and carry on confidently without alarming the patient was a much-needed experience.” (Male student, Clinic B, 2024)

‘Collaborative Practice’ was also highlighted by students, as they reflected about the supervising dentists and peers:

> “Dr. XXX’s expertise and compassionate care were truly inspiring. Watching her navigate sensitive cases with professionalism and empathy reinforced the importance of working together to achieve the best outcomes for patients.” (Male student, Clinic D, 2025)

> “Dr. XXX led us through his approach to care and encouraged collaboration among students, sharing valuable insights into how he practices certain procedures.” (Female student, Clinic A, 2023)

> “This collaborative experience sparked my interest in practicing in this community, even if only a few days a month. The staff and dentist were extremely supportive and made our work easier through collaboration.” (Male student, Clinic B, 2023)

The reflections also emphasized ‘Enactive Learning’, particularly regarding learning directly from supervising dentists:

> “I am very grateful to gain more experience in endodontics, as root canals are hard to come by at the university clinic. It was a great learning experience to try a new endodontic system and see how it works.” (Female student, Clinic B, 2022)

> “Dr. XXX shared many tips and clinical insights between patients, some of which I applied during later extractions, including the use of surgical instruments not available at the university clinic.” (Male student, Clinic A, 2023)

From the theme **In Spite of Differences**, students reflected on ‘Operational Procedures’ and variations among clinics:

> “The clinic’s setup differed significantly from the university clinic, particularly the software used for charting. While it required quick adjustment, I learned that this software is commonly used in private practice.” (Female student, Clinic B, 2024)

> “What surprised me most was how normal everything felt, despite the clinic functioning differently. It reminded me to keep my biases in check and remain open-minded moving forward in my career.” (Male student, Clinic D, 2023)

Students also discussed ‘Constrained Resources’, emphasizing adaptability and flexibility:

> “This rotation taught me how to work efficiently with different materials and instruments and underscored the importance of flexibility and resourcefulness in any clinical setting.” (Male student, Clinic A, 2024)

> “One major difference was the lack of composite shade options. At UBC we are spoiled with choices, but here we selected the darkest available shade. The patient was satisfied despite the imperfect match.” (Female student, Clinic B, 2023)

> “Differences in workflow, such as recall exams at the hygiene chair and alternative extraction instruments, highlighted how clinical setups vary based on available resources.” (Male student, Clinic D, 2023)

As the 4^th^ theme, **Pause–Breathe–Refine** emerged as a broader lesson for both students and academic institutions. One category, ‘Vicarious Quandary’, reflected students’ frustration with limited hands-on experience:

> “While grateful for the opportunity to observe and assist, I wished for more hands-on experience, as the principal dentist performed most procedures.” (Female student, Clinic A, 2022)

> “We are about to graduate, yet we are still observing? Spending the whole day on the sidelines was not productive. Even discussing cases or engaging more actively would have been beneficial.” (Male student, Clinic C, 2025)

> “I felt disappointed that the dentist took over the only procedure of my day, leaving me with little opportunity to practice.” (Female student, Clinic D, 2024)

Another category, ‘Foreigner Environment’, captured how unfamiliar and disorganized some sites felt to students:

> “The experience was overwhelming. We were expected to manage everything from setup to reprocessing without proper orientation to the clinic’s layout or technology.” (Female student, Clinic D, 2022)

> “Upon arrival, I was informed the rotation had been cancelled due to lack of a dentist, but we had never been notified.” (Male student, Clinic A, 2024)

> “This experience taught me that outside the university clinic, dentistry does not always go according to plan, and compromises must sometimes be made based on patient preferences.” (Female student, Clinic B, 2023)

## Discussion

Our exploratory thematic study revealed moments of revelation and struggles described by senior dental students reflecting on CSL placements serving equity-deserving communities at UBC’s Faculty of Dentistry. Consistent with existing literature, most reflections conveyed positive and transformative learning experiences. Themes related to professional growth and collaborative practice highlighted the potential influence of CSL on students’ willingness to work with underserved populations after graduation. Students also emphasized gains in independence and adaptability within community-tailored care, reinforcing prior findings that community placements provide learning opportunities not readily available in university clinics as discussed by Taylor, Carr, and Kujan (2024)^3^ and others.^4,21^

Students also discussed their views that they are addressing the unmet needs of equity-deserving groups.^6,22,23^ Such views were echoed in many reflections, particularly when students considered the severity of the cases they treated. Unmet needs were also emphasized in relation to communication strategies for patients with limited English proficiency, as discussed by Ho and colleagues (2024).^24^ These strategies are especially relevant in multicultural societies where English may not be spoken fluently, including Canada.^25,26^

Within the “financial abyss” category, students frequently highlighted the financial constraints faced by community members. Affordability is a key determinant of access to oral health care in jurisdictions where dentistry is primarily a private enterprise.^27,28^ It remains to be seen whether the newly introduced Canadian Dental Care Plan (CDCP), intended to make dental care more affordable for the majority of eligible Canadians receiving care in community clinics, will adequately address oral health inequities.^29^ The reflections underscore the role of social determinants in oral health inequities.

Students value and enjoy opportunities outside university clinics as discussed in prior research.^2,4,27^ These perspectives counter the reluctance of some training institutions to place students beyond the controlled university environment, which typically offers abundant faculty oversight, standardized equipment, and predictable scheduling.^5^ The exposure to alternative clinical settings may provide students with a more realistic understanding of the profession and better prepare them for real-world practice.

Similarly, the opportunity to learn from more experienced dental providers in diverse settings was echoed in many reflections categorized under “constrained resources,” “operational procedures,” “enactive learning,” and “collaborative practice.” Soon-to-be junior dentists clearly benefit from role models who play creative and influential roles in fostering professionalism and shaping career trajectories.^30,31^

Despite the positive learning experiences, students highlighted the detrimental impact of, for example, overly controlling mentorship styles and observation-only experiences on their learning. That is, mentorship styles that are overly controlling, disengaged, or disempowering should be avoided in health care education broadly,^32^ and in dentistry in particular.^33^ Students identified such concerns under the theme ‘vicarious quandary.’ One of the main implementation challenges of clinical placements relates to instructor calibration, as recently discussed by Dana and colleagues (2025).^13^ Additional challenges identified in both the literature and students’ reflections included logistical barriers (e.g., patient cancellations, variability in clinic infrastructure) and the need to adequately prepare students for unfamiliar environments, including clinic operations and patient populations.^14^

When considering types of experiences, clinical placements that are solely observational may be better positioned earlier in the curriculum, unless they are intentionally designed to alternate between observation and hands-on practice, as suggested by Horst et al.^34^ and others.^35^ To address these challenges, CSL activities should be thoughtfully designed, with clearly articulated learning objectives and outcomes, comprehensive onboarding processes, and well-calibrated instructors. Student expectations must align with their level of training, and partnerships between academic institutions and community sites should be collaborative and mutually beneficial to maximize outcomes for both learners and patients.^5,14^

Although informative, our study has several limitations. Because the data were drawn from a single institution, albeit across multiple student cohorts and community placements, generalizability to other schools and programs is limited. Although the reflections were not graded, they may still be subject to social desirability bias, as students may have emphasized more positive aspects of their experiences. Nevertheless, many students also shared critical perspectives and lessons learned, which strengthens the study’s contribution. Future research should examine CSL clinical activities after the implementation challenges identified here have been addressed. Additionally, studies exploring community members’ perspectives on student involvement and the care they receive are warranted.

## Conclusions

The CSL placements described by senior dental students were meaningful, educational, and transformative. Their reflections highlight the importance of community-based clinical education for professional growth, social awareness, and service to equity-deserving populations. Implementation challenges were also identified, including limited hands-on opportunities, inconsistent supervision, logistical disruptions, and insufficient orientation to unfamiliar clinical environments. These findings align with broader concerns regarding instructor calibration, student preparedness, and alignment of learning objectives in CSL programs. Intentional program design, comprehensive onboarding, instructor calibration, and alignment of expectations with students’ level of training are essential to maximizing the educational and service value of CSL. Strong, reciprocal partnerships between academic institutions and community sites remain critical. These findings support the continued integration of CSL into dental curricula and underscore the need to address implementation challenges to optimize learning outcomes.

## Data Availability

De-identified data produced in the present study may be available upon reasonable request to the authors

## Authors’ contributions

Dr Brondani contributed to the conception of this manuscript, design and acquisition of the literature, as well as drafting and critically revising the manuscript; Dr Pereira and Ms Elias contributed to the thematic analysis and interpretation of the textual data as well as with the literature review. The authors disclose the use of ChatGPT (https://chat.openai.com/) exclusively for language editing, including grammar, formatting, and clarity. No artificial intelligence–generated content (AIGC) tools or other large language models (LLMs) were used in the development of the manuscript’s content.

## Acknowledgments

The community clinics and hospital sites are acknowledged for supporting student learning and care delivery. We are grateful to the students whose reflections made this study possible. The authors declare no conflicts of interest and confirm accountability for all aspects of the work. The authors acknowledge the use of ChatGPT (https://chat.openai.com/) to improve grammar, formatting, and clarity, while taking full responsibility for the content and thematic analyses.

## References

1 Jessani A, Athanasakos A, Peltz R, Hussain R, Radhaa A, McIntosh M, Lathif A, McLean S. Training Socially-Conscious Dentists: Development and Integration of Community Service-Learning in Dental Curricula in Ontario, Canada. International Dent J. 2025;75(3): 1874–1884,

2 Brondani MA, Clark C, Rossoff L, Aleksejūnienė J. An evolving community based dental course on Professionalism and Community Service. J Dent Educ. 2008; 72: 1160–1168.

3 Taylor M, Carr S, Kujan O. Community-Based Dental Education (CBDE): A Survey of Current Program Implementation at Australian Dental Schools. Int J Dent. 2024:2890518.

4 Bahammam, H.A., Bahammam, S.A. Service-learning’s impact on dental students’ attitude to community service. BMC Med Educ 2023; 23: 59.

5 Brondani M, Dawson AB, Jessani A, Donnelly L. The fear of letting go and the Ivory Tower of dental educational training. J Dent Educ. 2023; 87(11):1594–1597.

6 Mays KA, Maguire M. Care Provided by Students in Community-Based Dental Education: Helping Meet Oral Health Needs in Underserved Communities. J Dent Educ. 2018;82(1):20–28.

7 Shukla A, Amrutham V, Albright A. Comparison of clinical independence level scores among predoctoral dental students between dental school clinic and community clinic rotation. J Dent Educ. 2025;89(7):1106–1116.

8 Campbell F, Rogers H. Through the looking glass: a review of the literature surrounding reflective practice in dentistry. Br Dent J. 2022;232(10):729–734.

9 Brondani M. Students’ reflective learning within a community-service learning dental module. J Dent Educ 2010; 74: 628–636

10 Brondani M, Harjani M, Siarkowski M, Adeniyi A, Butler K, Dakelth S, Maynard R, Ross K, O’Dwyer C, Donnelly L. Community as the teacher on issues of social responsibility, substance use, and queer health in dental education. PLoS One. 2020;15(8):e0237327.

11 Brondani MA. Teaching social responsibility through community service-learning in predoctoral dental education. J Dent Educ. 2012;76(5):609–19.

12 Deogade SC, Naitam D. Reflective learning in community-based dental education. Educ Health (Abingdon). 2016;29(2):119–23.

13 Dana E, Fitzgerald M, Kinney J, Riaz M, Cullen J. Comparison of community-based clinical education implementation among US dental and dental hygiene education programs. J Dent Educ. 2025;89(6):922–932

14 Ayibuof u-Uwandi V, Dyer TA. What are the public health benefits of community-based education in dentistry? A scoping review. Br Dent J. 2024 doi: 10.1038/s41415-024-7908-4.

15 Brondani M, Harjani M, Alfazwzan N, et al. Discussing elder abuse and neglect in undergraduate dental education: a commentary. J Elder Abuse Negl 2020; 32(4): 399–408.

16 Braun V, Clarke V, Anderson S, et al. Thematic Analysis: A Practical Guide. London, United Kingdom: SAGE Publications; 2021. p. 376.

17 Brondani MA, Bryant SR, MacEntee MI. Elders’ assessment of an evolving model of oral health. Gerodontology 2007; 24: 189–195.

18 Brondani M, Donnelly L. COVID-19 pandemic: Students’ perspectives on dental geriatric care and education. J Dent Educ 2020; 84(11): 1237–1244.

19 Gale NK, Heath G, Cameron E, Rashid S, Redwood S. Using the framework method for the analysis of qualitative data in multi-disciplinary health research. BMC Med Res Methodol. 2013;13:117.

20 Purssell E, Gould D. Undertaking qualitative reviews in nursing and education - A method of thematic analysis for students and clinicians. Int J Nurs Stud Adv. 2021;3:100036.

21 Taylor MR, Carr SE, Baynes L, Kujan O. Student and clinical supervisor perceptions of community-based dental educational experiences: A scoping review. J Dent Educ. 2024;88(6):798–814.

22 Mofidi M, Strauss R, Pitner LL, Sandler ES. Dental students’ reflections on their community-based experiences: the use of critical incidents. J Dent Educ. 2003;67(5):515–23

23 Brondani MA, Chen A, Chiu A, Gooch S, Ko K, Lee K, Maskan A, Steed B. Undergraduate geriatric education through community service learning. Gerodontology. 2012;29(2):e1222–9.

24 Ho JCY, Chai HH, Lo ECM, Huang MZ, Chu CH. Strategies for Effective Dentist-Patient Communication: A Literature Review. Patient Prefer Adherence. 2024;18:1385–1394.

25 Doucette H, Yang S, Spina M. The impact of culture on new Asian immigrants’ access to oral health care: a scoping review. Can J Dent Hyg. 2023;57(1):33–43

26 Zanchetta MS, Poureslami IM. Health literacy within the reality of immigrants’ culture and language. Can J Public Health. 2006; 97 Suppl 2:S26–30

27 Thompson B, Cooney P, Lawrence H, Ravaghi V, Quiñonez C. The potential oral health impact of cost barriers to dental care: findings from a Canadian population-based study. BMC Oral Health. 2014;14:78

28 Heaton LJ, Smith TA, Raybould TP. Factors influencing use of dental services in rural and urban communities: considerations for practitioners in underserved areas. J Dent Educ. 2004;68(10):1081–9.

29 Brondani M, Jessani A, Faria-e-Silva A, Ardenghi D. Delivering oral health care in Canada under the new Canadian Dental Care Plan: initial findings. Journal of the Canadian Dental Association 2025, 91:p15

30 Modha B. Experiential learning without prior vicarious learning: an insight from the primary dental care setting. Educ Prim Care. 2021; 32(1):49–55.

31 Osama OM, Gallagher JE. Role models and professional development in dentistry: an important resource: The views of early career stage dentists at one academic health science centre in England. Eur J Dent Educ. 2018;22(1):e81–e87.

32 Keinänen AL, Lähdesmäki R, Juntunen J, Tuomikoski AM, Kääriäinen M, Mikkonen K, Effectiveness of mentoring education on health care professionalś mentoring competence: A systematic review. Nurse Educ Today. 2023; 121: 105709

33 Nathwani S, Rahman N. GROWing in dentistry: mentoring the dental professional. Br Dent J. 2022;232(4):261–266.

34 Horst JA, Clark MD, Lee AH. Observation, assisting, apprenticeship: cycles of visual and kinesthetic learning in dental education. J Dent Educ. 2009;73(8):919–33

35 Hasanzade M, Ghazanfari R, Siadat H, Kharazifard MJ. Effect of Clinical Rotation Timing on Learning Quality of Dental Students. Front Dent. 2022;19:33.

